# Demographic Representation of a Decentralized Long COVID Trial: A Comparison with the 2023 National Health Interview Survey

**DOI:** 10.1101/2024.11.25.24317941

**Authors:** Mitsuaki Sawano, Frederick Warner, Yashira Henriquez, Bornali Bhattacharjee, Akiko Iwasaki, Harlan M. Krumholz

## Abstract

**Importance:** Clinical trials can be a chokepoint to progress in medicine. They are commonly expensive, slow, and not participant-centric. Ensuring patient-centric, equitable trials with diverse demographic representation is essential for developing generalizable treatments and reducing healthcare disparities. Thus, there is an urgent need to test new ways of conducting clinical trials. The PAX LC trial is a novel Phase II decentralized, digital, and participant-centric double-blind randomized trial of Paxlovid for long COVID and is a test case for a new approach. There is a need to determine how well the trial enrolled a representative population with long COVID.

**Objective:** To compare the demographic characteristics of participants in the PAX LC trial with those reported in the National Health Interview Survey (NHIS) to assess how well the decentralized recruitment reflects the broader population in the US with long COVID.

**Design:** Descriptive analysis comparing 2 databases: a Phase 2, 1:1 randomized, double-blind, superiority, placebo-controlled trial conducted in community-dwelling adults with long COVID, and a nationwide observational cross-sectional survey.

**Setting:** Decentralized clinical trial with participants recruited from 48 contiguous states in the US. The NHIS survey participants were recruited from all 50 states.

**Participants:** 100 highly symptomatic adults with long COVID in the PAX LC trial, compared with 575 from NHIS.

**Intervention(s) or Exposure(s):** None.

**Main Outcome(s) and Measure(s):** Percentage distribution by age group, gender, race and ethnicity, and region for each group.

**Results:** PAX LC included 100 participants and NHIS 575, with PAX LC participants being younger (median 42 vs. 53 years). There was an equal proportion of women. Non-Hispanic White representation was higher in PAX LC (89% vs. 61% in NHIS), while the Hispanic and Non-Hispanic Black groups were underrepresented (5% vs. 20% and 1% vs. 12%, respectively). More PAX LC participants were from the Northeast (51% vs. 15%) due to earlier enrollment in this region.

**Conclusions and Relevance:** The PAX LC trial highlights the potential of decentralized trials to achieve geographic diversity and balanced age and gender representation. However, to maximize the benefits of this approach, further efforts are warranted to improve racial and ethnic diversity.

**Trial Registration:** NCT05668091.

## INTRODUCTION

Clinical trials can be a chokepoint to progress in medicine. They are commonly expensive, slow, and not participant-centric. Ensuring patient-centric, equitable trials with diverse demographic representation is essential for developing generalizable treatments and reducing healthcare disparities. Thus, there is an urgent need to test new ways of conducting clinical trials.

Decentralized clinical trials offer a potential solution to this issue by increasing accessibility for participants who may otherwise be unable to engage in traditional trials due to geographic, socioeconomic, or healthcare access barriers. By leveraging technology, remote data collection, and flexible trial designs, decentralized trials can help ensure more inclusive recruitment across diverse populations. However, the decentralized design may also adversely affect the representativeness of the study population.

The PAX LC trial (An Interventional Decentralized Phase 2, Randomised, Double-Blind, 2-Arm Study to Investigate the Efficacy and Safety of Orally Administered Nirmatrelvir/Ritonavir Compared With Placebo/Ritonavir in Participants With Long COVID: NCT05668091) was designed to improve trial access and speed enrollment with high efficiency and participant-centricity. It was a Phase 2, 1:1 randomized, double-blind, superiority, placebo-controlled trial in 100 community-dwelling, highly symptomatic adult participants with long COVID residing in the 48 contiguous US states to determine the efficacy, safety, and tolerability of 15 days of nirmatrelvir/ritonavir compared with placebo/ritonavir.^1^

This study compared the demographics of PAX LC trial participants with those from the nationally representative, population-based National Health Interview Survey (NHIS) conducted in 2023. The goal was to assess how effectively the decentralized recruitment produced a representative sample regarding gender and racial or ethnic diversity.

## METHODS

We compared the demographic characteristics of participants in PAX LC with those of the 2023 NHIS. PAX LC participants were recruited between April 2023 and April 2024, while the NHIS participants were recruited in the year 2023. The PAX LC trial recruited participants from 48 contiguous

US states, whereas the NHIS recruited participants from all 50 states. Recruitment for PAX LC began in 3 Northeast states and gradually expanded to 44 states by August 2023, with eventual coverage of all 48 contiguous states by November 2023. Participants could join the study from various channels, including the study website, social media posts, podcasts, webinars, Yale press releases, Yale Help Us Discover volunteer database, and Yale Cultural Ambassadors.^2-7^ The study also had a preset diversity goal of recruiting 20% from underrepresented populations.

The PAX LC trial used the World Health Organization’s definition of long COVID: Long COVID occurs in individuals with a history of probable or confirmed SARS-CoV-2 infection, usually 3 months from the onset of COVID-19 with symptoms that last 2 months and cannot be explained by an alternative diagnosis. For this trial, this definition is operationalized as prior infection with SARS-CoV-2, as documented in the participant’s medical record, followed by the onset of symptoms within 4 weeks of the index infection that persist for at least 3 months after the index infection. Participants were required to have a baseline general health status rated as “fair” or “worse,” with a self-reported “good” or better health status before their index infection and no other evident reason for the decline in general health.

To enable a meaningful comparison with the 2023 NHIS, we limited the sample to individuals who had either a confirmed positive COVID-19 test or a doctor’s diagnosis of COVID-19, responded “yes” to the question, “Did you have any symptoms lasting 3 months or longer that you did not have before having coronavirus or COVID-19?”, and also rated their current health as “fair” or “poor” in response to the question, “Would you say your health in general is excellent, very good, good, fair, or poor?” We further excluded individuals residing in Alaska, Hawaii, Guam, Puerto Rico, and the Virgin Islands, since the PAX LC trial was only available in the 48 contiguous states.

The median and interquartile ranges for age were calculated for the 2 cohorts. In the NHIS dataset, 17 people over 85 years of age were suppressed in the publicly available dataset and were counted as 85 years of age in the current analysis. Furthermore, sample weights were used to calculate the proportion of participants in each categorical variable in the NHIS. All statistical analyses were performed using R, version 4.1.2 (R Foundation for Statistical Computing). The Yale University Institutional Review Board approved the PAX LC study, and the use of NHIS data was exempt from approval, as both datasets are publicly available at the Centers for Disease Control and Prevention website.

## RESULTS

Overall, there were 100 participants in PAX LC from 28 states and 575 participants in the NHIS with long COVID from 50 states and the District of Columbia (Table 1 and Figure 1).

**Table 1.** Demographic Comparison of PAX LC and NHIS.

|  | **PAX LC (N = 100)** | **NHIS\* (N = 575)** |
| --- | --- | --- |
| **Age, median (IQR)** | 42 (34, 50) | 53 (39, 65) |
| **Female, Sex (%)** | 66% | 66% |
| **Race and ethnicity (%)** |  |  |
| Hispanic | 5.0% | 20% |
| Non-Hispanic AIAN | 0.0% | 1.3% |
| Non-Hispanic Asian | 4.0% | 1.8% |
| Non-Hispanic Black | 1.0% | 12% |
| Non-Hispanic Other | 1.0% | 3.3% |
| Non-Hispanic White | 89% | 61% |
| **Region (%)** |  |  |
| Midwest | 19% | 22% |
| Northeast | 51% | 15% |
| South | 11% | 41% |
| West | 19% | 22% |

\*Of the 575 people in the NHIS, 17 were labeled as 85 years of age due to suppression of small sample size categories.
AIAN: American Indian or Alaska Native; BRFSS: Behavioral Risk Factor Surveillance System; IQR: Interquartile Range; NHIS: National Health Interview Survey; PAX LC: An Interventional Decentralized Phase 2, Randomised, Double-Blind, 2-Arm Study to Investigate the Efficacy and Safety of Orally Administered Nirmatrelvir/Ritonavir Compared With Placebo/Ritonavir in Participants With Long COVID

**Figure 1.**
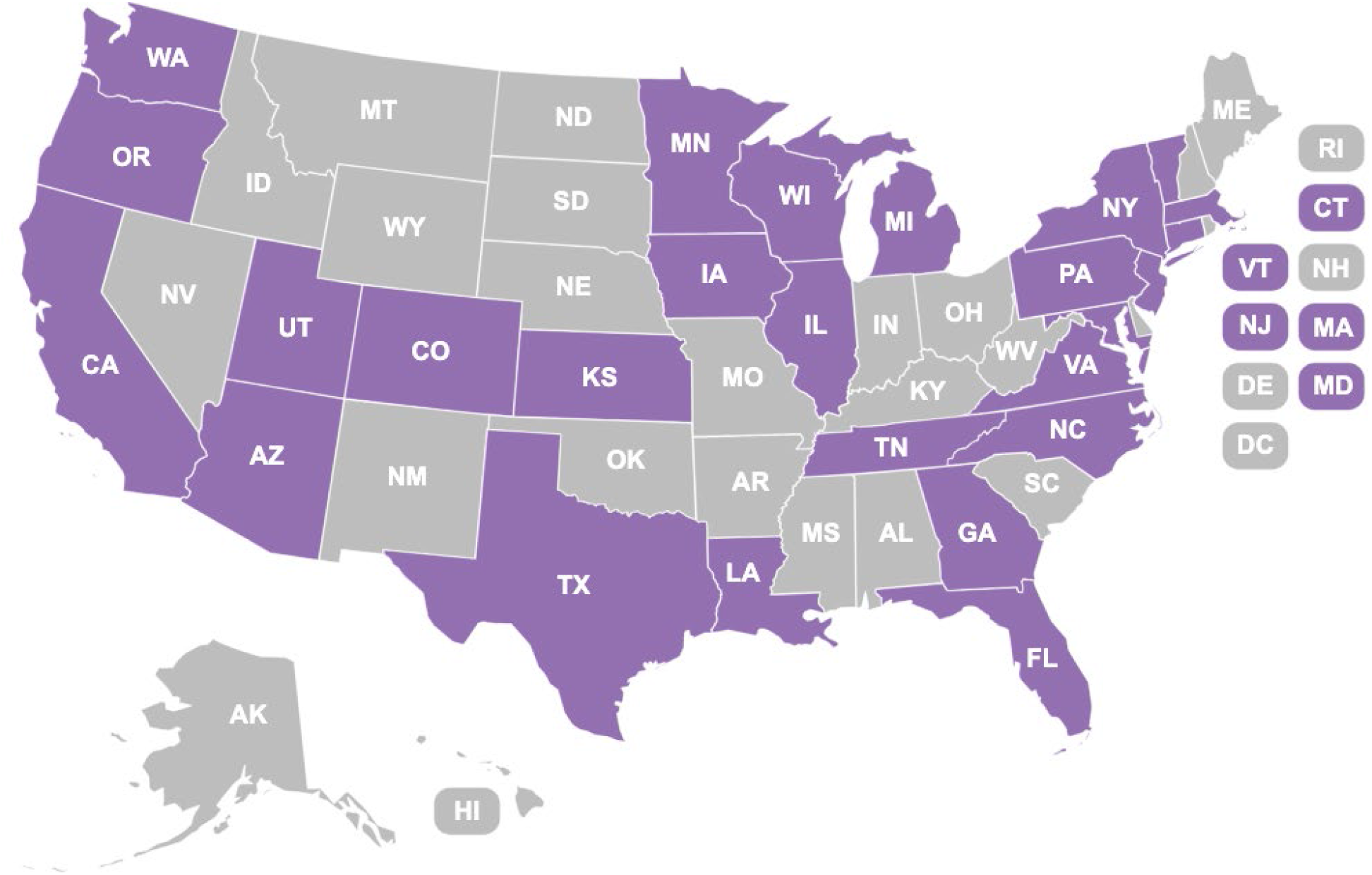
States with PAX LC trial participants. Map of the United States showing states with at least 1 participant enrolled in the PAX LC trial. States with participants are shaded in purple and states without any participants are unshaded.

The age distribution was younger in PAX LC, with a median age of 42 years (IQR 34–50) compared with 53 years (IQR 39–65) in NHIS. The sex distribution was similar across both groups, with approximately 66% identifying as female. PAX LC had a higher proportion of Non-Hispanic White participants (89%) than NHIS (61%). In contrast, other racial and ethnic groups, including Hispanic (5% in PAX LC vs. 20% in NHIS) and Non-Hispanic Black (1% in PAX LC vs. 12% in NHIS), were underrepresented. Participants in PAX LC were more likely to be from the Northeast (51%) than NHIS participants (15%) due to regional enrollment before the trial expanded nationally.

## DISCUSSION

The PAX LC trial enrolled a younger and more non-Hispanic White population than would have occurred had it been a nationally representative group. The sex distribution, however, was quite similar. Better representation remains an aspiration, but this Phase II study is only the first effort at a national Phase II decentralized trial. Although there are opportunities for further optimization, PAX LC was successful in rapidly enrolling a highly physically and mentally impaired group in a trial under one Institutional Review Board and no physical sites for in-person visits or assessments.

There are several key points from this study. First, female participants were adequately represented, and the distribution across age groups closely mirrored the demographic characteristics identified in nationwide surveys. Furthermore, the study successfully enrolled participants from all 4 regions, including the most rural US state (Vermont), demonstrating its capacity to capture geographically diverse populations. The diverse outreach strategies enabled more than 1000 individuals to access the prescreening questions and participate over a year, with approximately one-third passing the prescreening stage, ultimately resulting in 100 participants in the trial.

Second, PAX LC had a higher proportion of White participants and lower representation of underrepresented minorities, which may be explained by several factors. While the study team tried to engage Black people using various strategies, increasing representation may require greater involvement from physicians and investigators who have established trust within these communities. This reflects a broader challenge in making decentralized trials more representative, as it runs counter to the strength of such trials, which aim to reduce the need for in-person interaction. Notably, recent successful clinical trials with high Black American participation have often relied on in-person recruitment at churches and barbershops.^8^

Our study has some limitations. First, the smaller sample size of PAX LC compared with the NHIS may limit the generalizability of the findings. Second, while decentralized trials improve accessibility, they may still favor participants with access to technology, skewing recruitment toward certain socioeconomic groups. Nonetheless, in the PAX LC trial, the requirement for participants to have a health record documenting treatment for long COVID, a connection to health data from their usual source of medical care, and to speak and write fluently in English may have significantly contributed to underrepresentation in certain demographics.

In conclusion, the PAX LC trial highlights the potential of decentralized trials to achieve geographic diversity and balanced age and gender representation. However, to maximize the benefits of this approach, further efforts are warranted to improve racial and ethnic diversity.

## Data Availability

Anonymised individual participant data and study documents can be requested from the corresponding author and will be made available 12 months after publication of this paper, subject to the approval of the Trial Steering Committee and funding to anonymise the data.

